# Sensory stimulation enhances phantom limb perception and movement decoding

**DOI:** 10.1101/2020.05.22.20109330

**Authors:** Luke E. Osborn, Keqin Ding, Mark A. Hays, Rohit Bose, Mark M. Iskarous, Andrei Dragomir, Zied Tayeb, György M. Lévay, Christopher L. Hunt, Gordon Cheng, Robert S. Armiger, Anastasios Bezerianos, Matthew S. Fifer, Nitish V. Thakor

**Author notes:** (L.E.O.); (N.V.T.). these authors contributed equally (Keqin Ding, Mark A. Hays).

## Abstract

**Objective:** A major challenge for controlling a prosthetic arm is communication between the device and the user’s phantom limb. We show the ability to enhance amputees’ phantom limb perception and improve movement decoding through targeted transcutaneous electrical nerve stimulation (tTENS).

**Approach:** Transcutaneous nerve stimulation experiments were performed with four amputee participants to map phantom limb perception. We measured myoelectric signals during phantom hand movements before and after amputees received sensory stimulation. Using electroencephalogram (EEG) monitoring, we measure the neural activity in sensorimotor regions during phantom movements and stimulation. In one participant, we also tracked sensory mapping over 2 years and movement decoding performance over 1 year.

**Main results:** Results show improvements in the amputees’ ability to perceive and move the phantom hand as a result of sensory stimulation, which leads to improved movement decoding. In the extended study with one amputee, we found that sensory mapping remains stable over 2 years. Remarkably, sensory stimulation improves within-day movement decoding while performance remains stable over 1 year. From the EEG, we observed cortical correlates of sensorimotor integration and increased motor-related neural activity as a result of enhanced phantom limb perception.

**Significance:** This work demonstrates that phantom limb perception influences prosthesis control and can benefit from targeted nerve stimulation. These findings have implications for improving prosthesis usability and function due to a heightened sense of the phantom hand.

## 1. Introduction

Sensory information, specifically touch and proprioception, are essential for palpating, exploring, and manipulating objects in our surroundings [1]. Through sensory feedback and errors in our sensory predictions, we develop sophisticated internal models of sensorimotor integration [2], and we continue to update and strengthen our internal sensorimotor models for controlling limb movement [3]. Recently, researchers showed that supplementary auditory feedback can help improve internal models and performance in myoelectric control of a virtual prosthesis by able-bodied subjects [4], further indicating the role of feedback in sensorimotor control loops.

For upper limb amputees, the sensorimotor loop is severely disrupted as a result of limb loss; however, perception of the phantom limb persists for many [5]. Researchers made profound breakthroughs in providing naturalistic tactile sensations back to amputees by stimulating peripheral nerves, both invasively [6–9] and noninvasively [10–12], in the residual limb. Sensory feedback can provide perceptions of pressure [6, 7], enable discrimination of textures [8], create perceptions of movement across the phantom hand [9], help in reducing phantom pain [13], and improve prosthesis use at home [14]. Biomimetic stimulation models can enhance naturalness of the tactile sensation [15], improve object manipulation [16], and be used to provide receptor specific information to enable sensations of pressure or pain [12]. Kinesthetic illusions of phantom hand movement have also been produced using skin vibration on amputees who had undergone targeted muscle reinnervation (TMR) surgery [17]. Despite these successes, there is an unanswered question about the effect enhancing phantom hand perception has on the internal sensorimotor models that control phantom hand movements. Specifically, it is unclear how phantom hand perception affects motor function and resulting activation of muscles in the residual limb. Pattern recognition techniques aim to create a natural and intuitive control strategy for upper limb amputees by decoding movement from electromyography (EMG) signals in the residual limb [18]. Recently, proportional control of multiple degrees of freedom was achieved with derived motor unit action potentials in TMR subjects [19] and direct control using surface EMG electrodes [20].

We postulate that an important component of myoelectric decoding is the ability to perceive and move the phantom hand. Neural signals measured by EEG after TMR suggest that more natural cortical representations of the missing limb can occur in the motor cortex as a result of the surgery [21]. Additionally, recent results show somatosensory neural representation of the phantom limb exists even decades after amputation [22]. It is also known that movement representations persist in the motor cortex even when an amputee cannot generate voluntarily movements with the phantom hand, indicating that the lack of phantom control is not equivalent to the loss of neural representation [23]. Furthermore, evidence suggests that despite classical ideas of cortical reorginization after limb amputation, phantom limb representation of motor commands and muscle synergies still persist in the primary motor cortex [24]. Interestingly, activation of neural activity from sensory feedback through electrical stimulation occurred in both somatosensory and premotor regions during evoked phantom limb sensations [25].

In this work, we hypothesize that providing sensory stimulation to amputees can modulate the sensorimotor loop and enhance phantom limb perception, improving the ability to decode phantom hand movements using EMG pattern recognition (Fig. 1). Our study presents a number of important observations. Firstly, we demonstrate that sensory stimulation improves perception of the phantom hand. Secondly, we show that changes in phantom hand perception affect the ability to control phantom movements and a prosthesis. Finally, using EEG signals we show that increased activation of sensorimotor regions occur both during and after sensory stimulation and phantom limb activation.

**Fig. 1.**
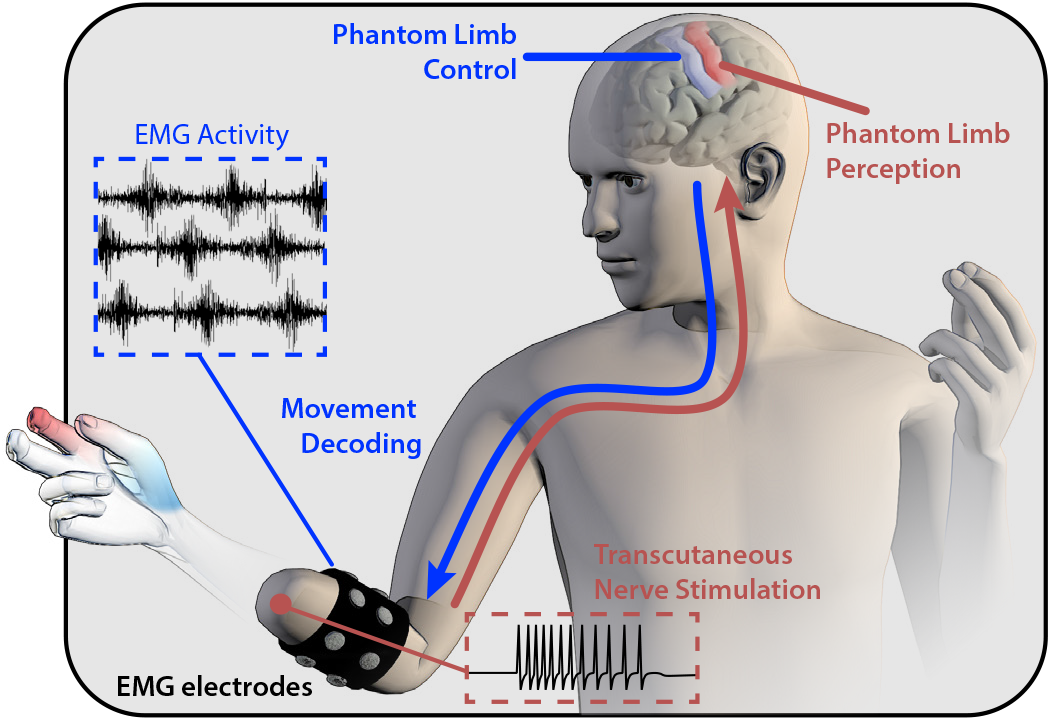
Phantom limb perception and control. Upper limb amputees often perceive their phantom limb. Voluntary movements of the phantom limb can be captured and decoded from electromyography (EMG) signals in the residual limb. We demonstrate the role of targeted transcutaneous electrical nerve stimulation (tTENS) to enhance phantom perception and improve movement decoding in a comprehensive investigation with 4 amputees.

## 2. Methods

Four male amputee participants with varying levels of prosthesis experience, ranging from none to over 8 yr, were recruited for this study and participated in at least one experiment (Table S1). Two amputee participants (A01 and A02) underwent elective amputations as a result of nerve injury, and three of the participants (A01, A03, and A04) have transhumeral amputations. Participant A02 has a transradial amputation. A03 also has a right arm transradial amputation but only uses a prosthesis on his left arm, which was the side used for the experiments in this study. Participants A01-A03 performed phantom hand movement tasks before and after receiving sensory stimulation to the phantom hand and took a user survey. A02 and A03 also participated in EEG recording experiments during phantom hand movements. A02 participated in a long-term study over 2 years to track changes in phantom sensory mapping and movement decoding performance. A04 participated in sensory mapping and the object movement task. Amputee participation is summarized in Table S1. All experiments were approved by the Johns Hopkins Medicine Institutional Review Boards. The amputees, who were recruited from previous studies or referrals, provided written informed consent to participate in the experiments.

### 2.1. Sensory stimulation

Sensory mapping was done with tTENS using a monopolar 1 mm beryllium copper (BeCu) probe connected to an isolated current stimulator (DS3, Digitimer Ltd., UK) to provide monophasic square wave pulses to underlying peripheral nerves, activating the phantom hand. This approach was validated in our previous work [12, 26]. An amplitude of 0.8 – 3.0 mA, frequency (*f*) of 2 – 4 Hz, and pulse width (pw) of 1 – 5 ms were used while mapping the phantom hand [12, 26]. Anatomical and ink markers were used, along with photographs of the amputee’s limbs, to map the areas of the residual limb to the phantom hand. For all other stimulation experiments, we used 5 mm disposable Ag/AgCl electrodes (Norotrode 20, Myotronics, USA) with *pw* = 1 ms and *f* = 45 Hz. Stimulation parameters were based on our previous work [12, 26] and were reliably detected by every participant.

### 2.2. EMG recording and movement decoding

For participants A01 and A02, 8 channels of raw EMG signals were measured using 13E200 Myobock amplifiers (Ottobock, Plymouth, MN) with bipolar Ag/AgCl electrodes placed uniformly around the circumference of the residual limb. No specific muscle groups were targeted for electrode placement. Signals were recorded by an NI USB-6009 (National Instruments, Austin, TX) at 1024 Hz with a 20 – 500 Hz digital bandpass and 60 Hz notch filters.

Participant A03 used a custom socket manufactured by his prosthetist (Dankmeyer, Linthicum, MD). Eight bipolar Ag/AgCl electrodes (Infinite Biomedical Technologies, Baltimore, MD) were embedded within the socket. The bipolar electrodes in the custom socket were amplified and filtered with a 20 – 500 Hz digital bandpass and 60 Hz digital notch filters.

EMG signal time domain features were extracted after 2 s of sustained movement using a 200 ms sliding window with new feature vectors computed every 50 ms. The features used were mean absolute value, waveform length, and variance (Supplementary Methods). Each movement cue was presented 3 times for 5 s and in a random order. Data from Day 187 of the long-term study was used for A03’s 9 class comparison because that was the first day he performed the movement decoding experiment before and after sensory stimulation. Movement decoding on Day 194 was done with 4 of the 8 electrodes in A03’s custom socket due to hardware failure. Data from 1 round of movements was discarded from each of A02’s visits due to hardware malfunction. For simultaneous tTENS with EMG recording from A02, grounding electrodes were placed on the residual limb to remove noise artifacts (Supplementary Methods, Fig. S1). Movements were decoded using the extracted EMG features with an LDA classifier. One-third of the EMG data was used as a holdout set from the training data for testing the classifier [18]. The classifier was trained and tested on data from the same day.

Participant A04 wore 2 EMG recording armbands (Myo, Thalmic Labs, CA), 8 stainless steel electrodes per band, on his residual limb, which was his typical setup for controlling the prosthesis used in this experiment during his daily activities. Filtered EMG data was collected from the armbands at 200 Hz. Training data was collected and movements were decoded using an LDA classifier on a custom controller embedded in the prosthesis [27].

### 2.3. EEG recording and analysis

Ag/AgCl EEG electrodes were used for recording neural activity at 500 Hz sampling frequency (64-ch, SynAmps2, Compumedics NeuroScan). Participant A02 and A03 took part in this experiment. Each participant was seated and tTENS electrodes were placed to activate the areas of the phantom hand corresponding to median, ulnar, and radial nerve innervation regions. Stimulation was for 2 s, followed by a 4 s delay with ± 25% time jitter before the next stimulation. The EEG data was band-pass filtered from 1 to 50 Hz and re-referenced to both mastoids. Automatic Artifact Removal (AAR) was used to remove the muscle (canonical correlation approach, 5 s window size) and ocular (blind source separation SOBI algorithm, 256 s window size) artifacts [28]. Independent component analysis (ICA) was used to remove additional artifacts. Continuous EEG data was epoched from 1 s before the start of each trial until 2 s after the stimulus presentation. All analysis was done using the EEGLAB toolbox in MATLAB [29].

The epoched EEG data was band-pass filtered from 8 Hz to 12 Hz to obtain the alpha band. We further epoched the data from 450 – 850 ms after the stimulus presentation to remove early activation due to tTENS and visual stimulation from analysis and focused on the motor-related activity in the brain. We evaluated the alpha band power relative to the total power of all bands in all electrodes for each condition and each trial. For each participant, phantom hand stimulation conditions (thumb and wrist for A02; thumb, pinky, and wrist for A03) were included for the rest of the analysis.

### 2.4. Experimental protocol

#### Phantom Movements with Stimulation

We used a modified Virtual Integration Environment (VIE) (Johns Hopkins University Applied Physics Lab (JHU/APL), Laurel, USA) in MATLAB to display movement cues. The subjects were seated in front of a screen that displayed the movement classes. The skin of the residual limb was cleaned with an alcohol wipe before tTENS and EMG electrode placement. The electrodes were allowed to settle for up to 10 min. After EMG data collection, the subject received tTENS. The sensory stimulation lasted between 30 – 60 min with continuous site activation for up to 10 s at a time. After tTENS, the participants performed another round of EMG data collection. Anatomical markers and photographs were used to ensure the electrodes were positioned in approximately the same location for A01 and A02. A03 used a customized socket, which ensured consistent electrode placement. The experiment lasted up to 3 hr.

For the long-term study, participant A03 performed periodic sensory mapping over 2 years (Day 1 - Day 738). A03 also performed 3 different phases of EMG data collection starting on Day 128 (labeled as Week 1 of the long-term EMG data). Fourteen movement classes were used (Fig. 5C-F, including a rest class). During Phase I (Week 1-6), A03 came in for an EMG recording session on average once per week. For Phase II (Week 8-10), he came in for EMG recording sessions on 4 different days. There were 3 separate rounds of EMG data collection on each of those days. EMG signals during the Pre-Stim condition were recorded for each movement and repeated 3 times. Next, movement cues were shown while sensory stimulation was being given to the phantom hand (Fig. 5). A final EMG recording session was performed without stimulation. There was up to a 30 min break between each of the 3 recording sessions. The total experiment lasted up to 3.5 hr each day. In Phase III (Week 12-48), A03 performed 4 follow-up EMG recording sessions. All EMG experiments were offline and participants didn’t receive feedback on EMG activity or decoding performance to prevent bias across the testing conditions.

#### Object Movement Task

Participant A04 used a modified VIE to interface with the pattern recognition software and prosthesis controller. The Modular Prosthetic Limb (MPL) [27], developed by JHU/APL, was mounted to the osseointegrated implant. A04 completed the object movement task before any sensory stimulation (Pre-Stim). A04 underwent tTENS sensory mapping and phantom hand activation for approximately 1.5 hr before completing the object movement task again after the sensory stimulation (Post-Stim). A new LDA pattern recognition classifier was trained before performing the object movement task for both the Pre-Stim and Post-Stim conditions. Movement classes used were hand open, tripod grasp, elbow (flexion and extension), and wrist (pronation and supination). Each movement class contained up to 5 s of training data. Each trial consisted of 5 repetitions of grabbing the object, moving it approximately 60 cm, and then releasing it. Participant A04 performed 3 trials of the task in both the Pre-Stim and Post-Stim conditions. No tactile feedback or tTENS was provided to A04 during the object movement task. Time to complete the task was recorded for each trial. The participant successfully moved the object every trial without dropping it.

#### Neural Recording

The participants were seated and shown visual movement cues with corresponding stimulation in median, ulnar, and radial regions, respectively (Fig. 5). Participant A02 was shown hand open and close. Participant A03 was shown tripod, index point, and wrist flexion. Baseline activity was recorded for up to 2 min. *Pre-Stim*: the participant mimicked movement cues with his phantom hand. *Stim*: the participant received tTENS to activate the phantom hand, but did not perform movements with his phantom hand. *Stim-Move:* the participant received sensory stimulation while performing movements with his phantom hand. *Post-Stim*: the participant performed phantom hand movements but with no sensory stimulation. For participant A02, each movement cue was presented 30 times for all conditions. For participant A03, each movement cue was presented 10 times for the Pre-Stim condition and 20 times for all other conditions.

For all experiments, results from data collected over multiple trials of the same experiment were averaged together.

Statistical *p* values were calculated using a two-tailed, two-sample *t* test and error bars represent the standard error of the mean, unless otherwise specified. All analysis was performed using MATLAB (MathWorks, Natick, USA).

## 3. Results

### 3.1. Sensory stimulation enhances phantom hand perception

For each participant, we used sensory mapping to identify regions of phantom hand activation. Targeted transcutaneous electrical nerve stimulation (tTENS) was used to activate underlying peripheral nerves in the residual limb, a method which was used in previous studies (Supplementary Discussion) [10, 12, 26]. Stimulation of the mapped regions on the residual limb resulted in perceived sensation in the phantom hand. Each amputee’s perception of their phantom limb is different and tTENS activated different phantom regions (Fig. 2). Sensations were reported primarily as tactile and included pressure, buzzing, vibration, and in the case of A01, a sensation of cold temperature on the palmar side of the middle and ring fingers (Fig. 2A).

**Fig. 2.**
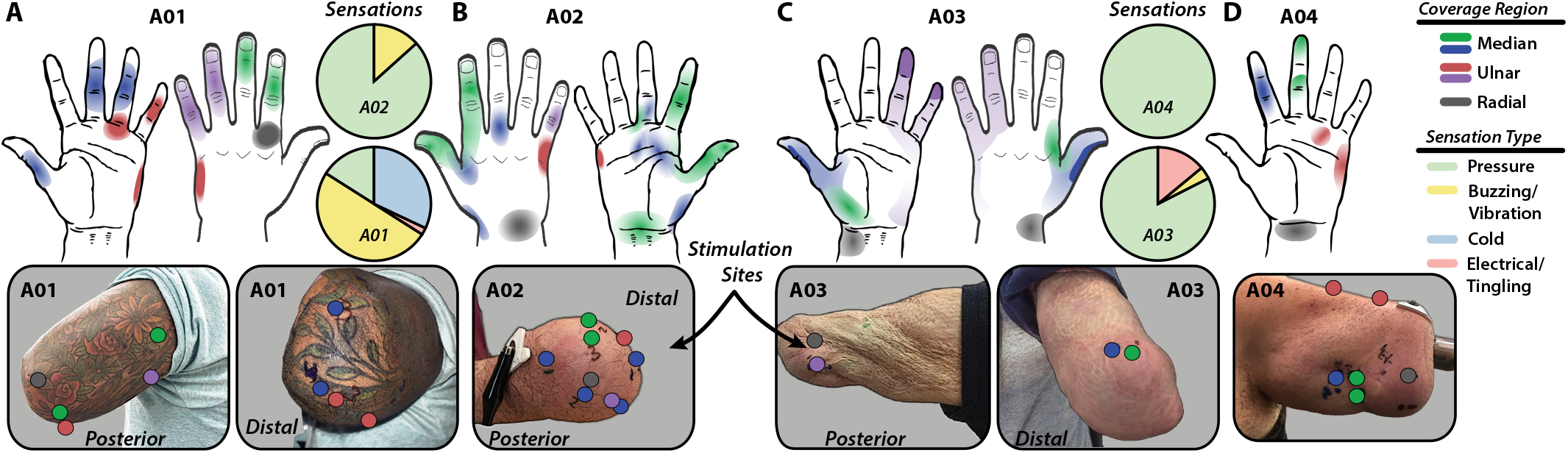
Sensory mapping of amputee participants. **(A)** Participant A01 reported sensations of general tactile activation, primarily buzzing or vibration, along with sensations of temperature changes on the palmar side of the middle and ring fingers. **(B)** Participant A02 reported sensations of pressure in the activated regions. The thumb and index finger, along with the ulnar and palmar sides of the hand, were the primary regions of activation. **(C)** Participant A03 perceived sensations of pressure and occasional tingling in the thumb, pinky, and wrist regions of his phantom hand. **(D)** Participant A04 perceived sensations as pressure in his phantom hand. For all phantom hand sensory maps, regions of strongest to faintest activation are indicated by a gradient of solid to faded color. In general, stimulation sites on the residual limb are <5 mm in diameter but are made larger here for illustration.

A user survey to gauge phantom hand perception, based on a previous study [30], was given to participants A01-A03 at the end of the day after a testing session (Fig. 3). In general, participants felt as if something was touching the phantom hand during the sensory stimulation. Furthermore, all participants who took the survey felt as if they could better perceive and, more importantly, move their phantom hand as a result of the nerve stimulation (Fig. 3).

**Fig. 3.**
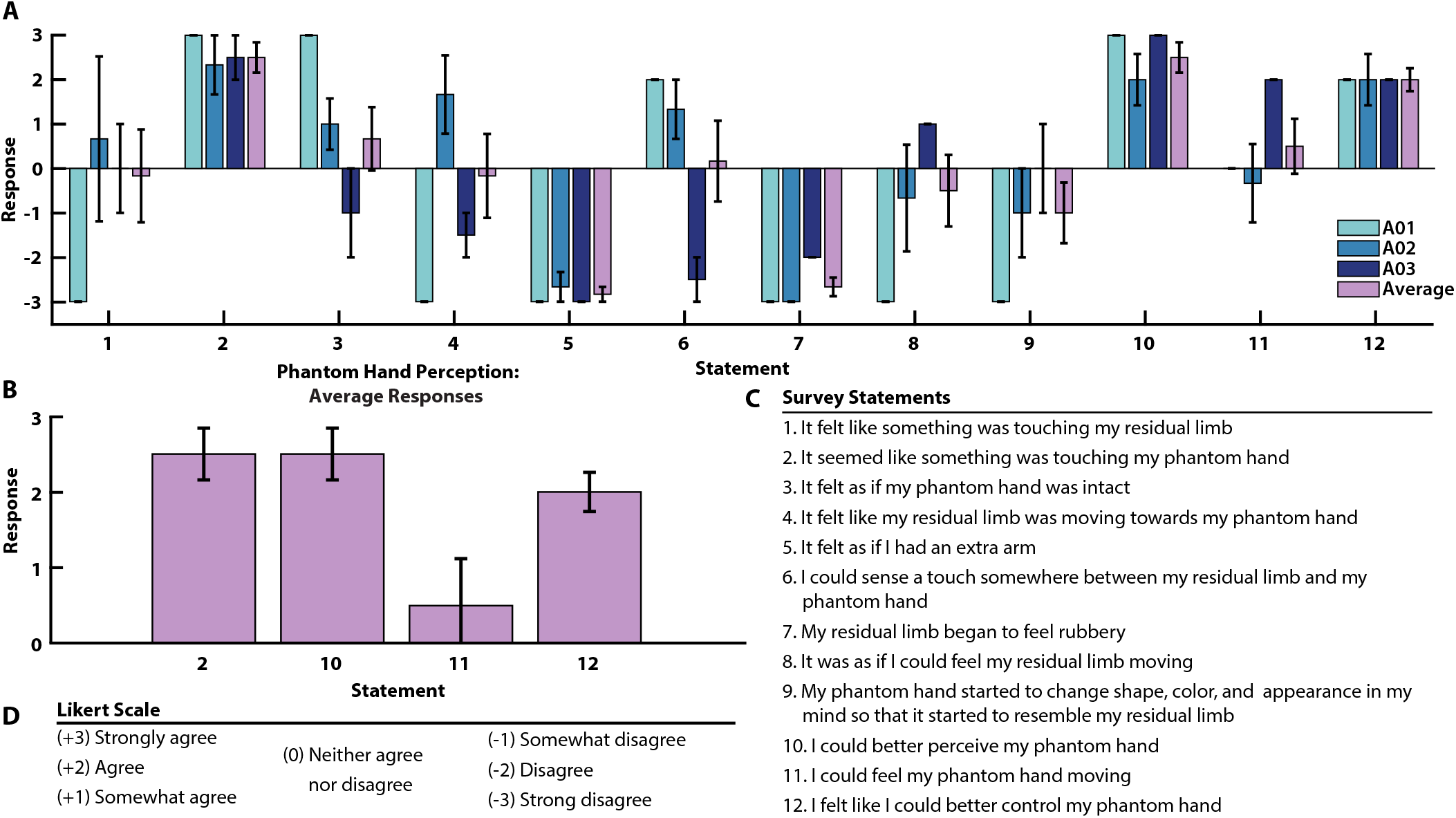
Sensory stimulation improves phantom perception as reported by user surveys. **(A)** User survey aimed at understanding subjective perception of sensory stimulation. Mixed results for several statements across participants suggest the varying nature of perception due to sensory stimulation. However, all participants agreed that heightened perception were a result of sensory stimulation through electrical nerve stimulation. Participants A02 and A03 took the survey twice on different days after sensory stimulation and phantom hand movement matching experiments. Participant A02 completed the survey again during a follow-up phone interview. The results were averaged. A01 took the survey once. **(B)** Averaged user results from survey response specifically on phantom hand perception as a result of sensory stimulation. For all participants, sensory stimulation enhanced perception of the phantom hand and importantly also giving the perception of better control over phantom hand movements. **(C)** Statements from the user survey. **(D)** The survey was scored using a Likert Scale with answers to statements ranging from “Strongly agree” (+3) to “Strongly disagree” (−3).

Participant A01 took the survey once, A02 completed the survey twice in person and an additional time during a follow-up phone interview, and A03 completed the survey twice. The survey was meant to gauge user perception of the phantom hand and sensory stimulation. All users reported enhanced perception and control of the phantom limb compared to normal baseline as a result of sensory stimulation. The statements were modeled after surveys from a previous study [30]. Results from the survey targeted specifically at quantifying the enhanced perception of the phantom limb as a result of sensory stimulation are shown in Fig. 3B. In general, participants felt as if something was touching the phantom hand during the sensory stimulation (Fig. 3A). Furthermore, all participants who took the survey felt as if they could better perceive and move (Q10 and Q12, respectively) their phantom hand as a result of the nerve stimulation (Fig. 3C). A04 did not take the survey, but he did verbally confirm that the sensory stimulation produced enhanced phantom hand perception. It should be noted that our survey does not capture changes in prosthesis embodiment or agency. The survey provides subjective responses from the participants to better understand their perceptions of phantom sensations.

### 3.2. Sensory stimulation improves movement decoding

Because sensory stimulation provides a heightened sense of the phantom hand (Fig. 3), we investigated the effect of this enhanced perception on the ability to make dexterous grasps with the phantom hand. Hand and wrist movements were visually presented to three of the participants (Fig. 4A, Fig. S2), who then attempted to mimic the movement with their phantom hand. Each amputee performed the hand and wrist movements before receiving any sensory stimulation (Pre-Stim). After EMG collection, regions of the phantom hand were activated via tTENS to provide general tactile sensation (see Methods).

**Fig. 4.**
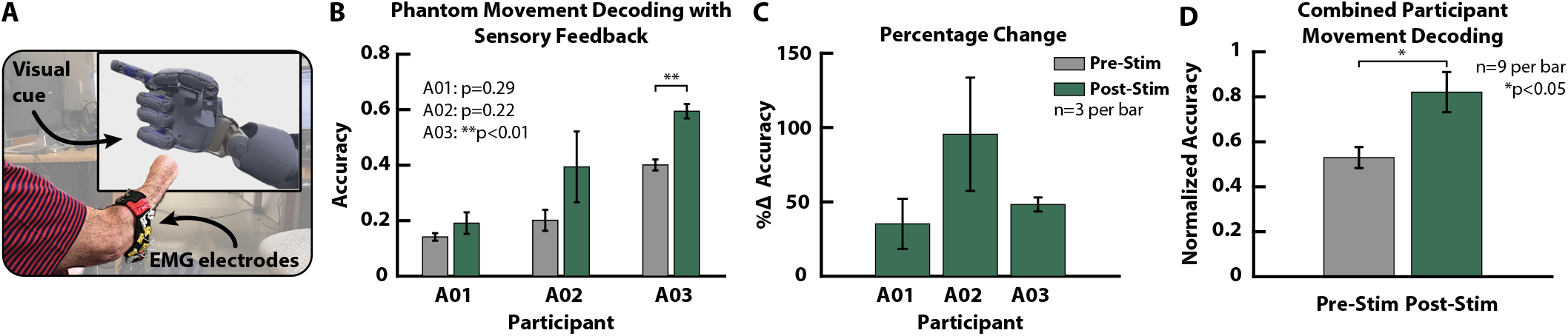
EMG performance of amputee subjects. **(A)** Five hand movements (rest, open, close, tripod, index point) and four wrist movements (pronation, supination, flexion, extension) were presented, one at a time, to the amputee participant, who attempted to match the movement with his phantom hand. **(B)** EMG decoding accuracy from the 9 movement classes before (Pre-Stim) and after (Post-Stim) sensory stimulation. **(C)** Percentage change in performance accuracy for all participants (absolute changes in Fig. S2). Relative performance increased at least 35% (A01) and up to 95% (A02) from baseline as a result of enhanced phantom limb perception. **(D)** The combined performance of all participants, normalized to the maximum individual performance, increased increased after sensory stimulation.

For participants A01-A03, the stimulation sites activated regions that covered the thumb, index, palm, and ulnar sides of the phantom hand (Fig. 2). The sensory stimulation session lasted up to 30 min and was followed by another round of EMG data collection (Post-Stim). The accuracy of the EMG movement classification is shown in Fig. 4B-D. Linear discriminant analysis (LDA), a standard EMG pattern recognition algorithm [18], was used to classify the movements. Results indicate at least a 35% increase in baseline EMG pattern recognition performance for all three participants (Fig. 4B-C). Improvements occur for all participants, but p<0.05 only in the case of A03. Overall, the averaged normalized decoding performance across all participants increased as a result of sensory stimulation (Fig. 4D).

### 3.3. Long-term sensory stimulation and EMG decoding

To better understand the influence of enhanced sensory perception on EMG pattern recognition performance, participant A03 took part in an extended study over 2 years. The primary regions of perceived activation were the thumb and index finger, the ulnar side, and the wrist of the phantom hand. These regions remained stable; that is, they did not migrate over the course of the study (Fig. 5A, Fig. S3). The structural similarity (SSIM) index [31] was calculated for each region across all the days and shows good similarity (>0.75) in all cases (Fig. 5B, Supplementary Methods). With every stimulation session, the participant verbally indicated an enhanced perception of his phantom hand during sensory stimulation. This subjective response was based on the daily baseline phantom hand perception before any stimulation experiments began.

**Fig. 5.**
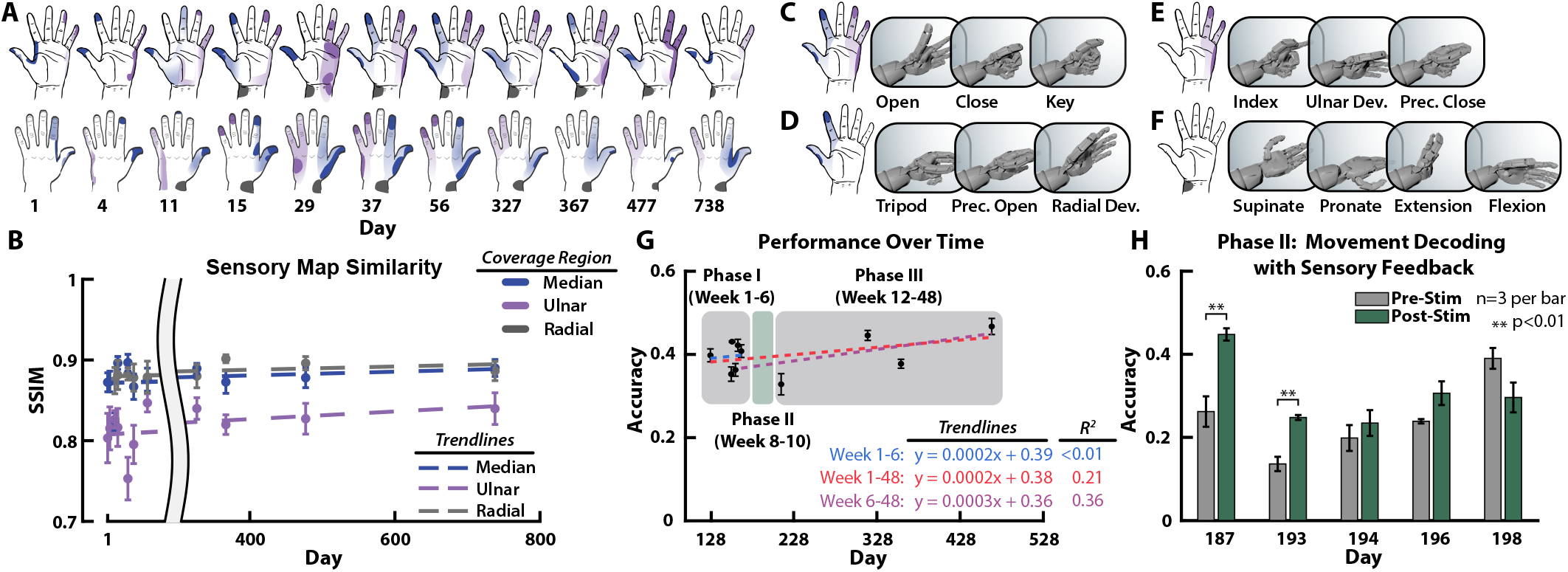
Long-term sensory mapping and movements. **(A)** Sensory mapping from tTENS of the ulnar, median, and radial regions was performed on participant A03 over a 2 year period. Activation maps of his phantom hand remained stable over the duration of the study with the primary regions of sensation being on his thumb and index finger, pinky, and wrist. **(B)** Structural similarity (SSIM) indices of sensory maps for each region show high similarity (>0.75) across the extended study. C-F, The participant associated activation of certain regions of his phantom hand to different grasp patterns. **(C)** Activation in the median and ulnar regions of his phantom hand were most closely associated with opening, closing, and the lateral key grasp. **(D)** Thumb and index finger, **(E)** ulnar, and **(F)** wrist region activations were associated with corresponding hand and wrist movements. No stimulation was provided during the rest class. **(G)** EMG pattern recognition performance was measured over nearly 1 year. An initial set of baseline data was collected in Phase I (Week 1-6), followed by a 3 week period of sensory stimulation through tTENS (Phase II, Week 8-10). Phase III (Week 12-48) consisted of sessions over a 37 week period. The subject was experienced with pattern recognition and showed a fairly consistent level of performance with a non-significant increase over time likely a result of continued prosthesis use (p>0.05, Fig. S4). **(H)** The stimulation phase shows improvements in EMG movement decoding of the 14 classes as a result of enhanced phantom limb perception (individual classes in Fig. S4). EMG signal recordings were taken for each movement class before (Pre-Stim) and after (Post-Stim) stimulation.

We investigated the effects of sensory stimulation on movement decoding performance compared to long-term performance over 1 year. The participant identified different regions of activation that best corresponded to particular movements of his phantom hand (Fig. 5C-F). The combinations of sensation in targeted regions of the phantom hand with movement classes were made based on what the amputee determined as being relevant phantom hand regions during attempted phantom movements. For example, during the index point and precision close hand movements the participant said he moved the thumb and index fingers but his main focus was on closing his pinky and ring fingers.

A custom prosthetic socket with embedded electrodes was used to ensure consistent electrode placement during each EMG recording session (Fig. S3). The long-term experiment was broken up into three phases. Phase I was 6 weeks long (Week 1-6) to establish a baseline in performance. Phase II (Week 8-10) was a 3 week period of sensory stimulation with EMG recordings. Phase III (Week 12-48) was a 37 week follow-up set of sessions to evaluate any lasting effects of the sensory feedback on the internal sensorimotor loop used by the amputee for moving his phantom hand (Fig. 5G). There were a total of 14 movement classes (Fig. 5C-F, 8 hand, including rest, and 6 wrist movements). During Phase II, EMG signals were recorded during each movement class before (Pre-Stim) and after (Post-Stim) stimulation. The EMG pattern recognition accuracy remained fairly stable for the 6 week period of Phase I with slightly more variation during Phase III. The sensory information provided to the phantom hand resulted in within-day improved movement decoding in most cases during Phase II (Fig. 5H) and with significant improvements in tripod and radial deviation movements; however, the effect did not appear to persist into Phase III (Fig. S4). The long-term changes during Phase III match the overall trend from the beginning of Phase I, indicating that short-term improvements from sensory reinforcement did not translate beyond individual days (Fig. 5G).

### 3.4. Movement decoding improves during sensory stimulation

We also investigated the effects of sensory stimulation during active movement. Participant A02 identified thumb and wrist areas of his phantom hand that, when activated, corresponded with specific movements (Fig. 6A). Movement decoding was performed on EMG signals recorded before (Pre-Stim), during (Stim), and after (Post-Stim) stimulation. Results show an obvious improvement in classifying attempted phantom hand movements during trials with sensory activation, whereas only a slight improvement is observed for trials after stimulation (Fig. 6B). The stimulation noise artifact was removed from the myoelectric signal using a hardware grounding approach (Supplementary Methods). A classwise comparison shows improvement in some movements during and after sensory stimulation but a decrease in others (Fig. S5).

**Fig. 6.**
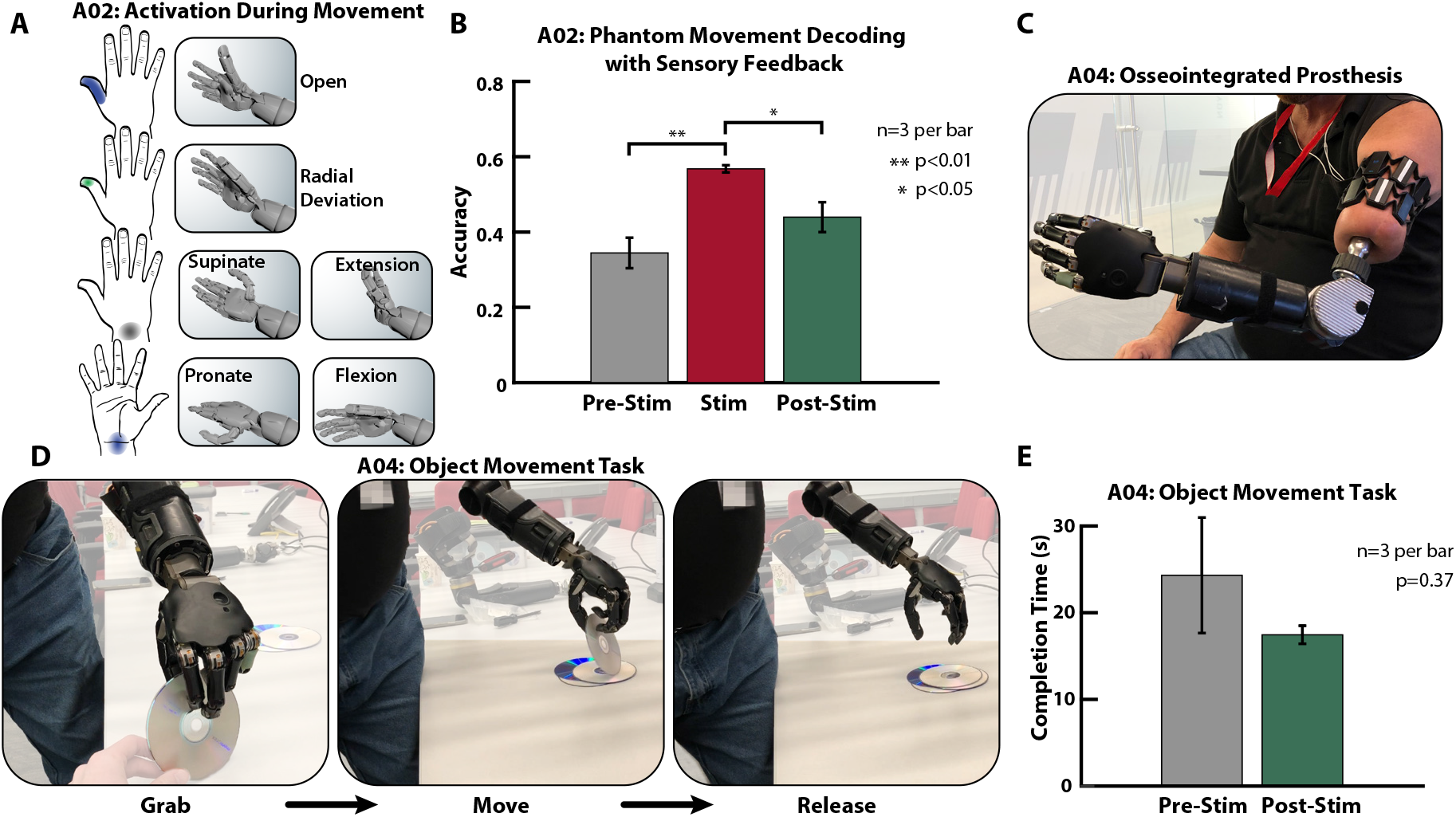
Movement decoding and prosthesis function improve after sensory stimulation. **(A)** A02 identified several wrist movements and corresponding phantom regions to receive sensation. **(B)** Movement decoding was done with EMG pattern recognition for trials before (Pre-Stim), during (Stim), and after (Post-Stim) stimulation to A02’s phantom hand. There was an increase in movement decoding during (Stim) and after (Post-Stim) stimulation, but the improvement is greater during the Stim condition. **(C)** Participant A04 used his osseointegrated prosthesis with EMG pattern recognition control for the functional task. **(D)** The object movement task consisted of grabbing, moving, and releasing an object using EMG pattern recognition prosthesis control. **(E)** The task completion time decreased after sensory activation of A04’s phantom hand (Post-Stim) as compared to the Pre-Stim task completion times.

### 3.5. Prosthesis control improves after sensory stimulation

We also tested the functional difference of an object grasping task with a prosthesis before and after sensory activation of the phantom hand in a fourth amputee participant. A04, who has an osseointegrated implant [32] and TMR, controlled a prosthesis using EMG pattern recognition (Fig. 6C). The participant underwent sensory mapping (Fig. 2d) and performed the object movement task before and after sensory activation of his phantom hand. The participant grabbed, moved, and released a compact disc using a tripod grasp (Fig. 6D). The average task completion time decreased in the Post-Stim condition (p=0.37, Fig. 6E). A04 did not take the user survey or participate in the EMG movement decoding experiments; however, the participant did verbally confirm that the sensory stimulation produced enhanced phantom hand perception.

### 3.6. Sensory stimulation increases EEG activity in sensorimotor regions

EEG signals were recorded to capture the neural activity in sensorimotor regions during sensory stimulation and phantom hand movement in participants A02 and A03. The alpha band (8 – 12 Hz) is relevant for sensorimotor-related activity [33, 34] and was used to evaluate the influence of sensory stimulation on phantom hand movement related neural activity.

The relative alpha power, the alpha power relative to the sum of power of all frequency bands, from the EEG was estimated for phantom hand movement before stimulation (Pre-Stim), during tTENS with no movement (Stim), during tTENS with movement (Stim-Move), and phantom hand movement after stimulation (Post-Stim) (Fig. 7A-D, G-J). Hand open and close were shown to participant A02 and tripod, index point, and wrist flexion movements were shown to participant A03. These classes were chosen by the participants because the classes were most closely associated with the tTENS phantom hand sensory mapping results. Furthermore, the classes and stimulation sites align with locations used by A03 in the long-term study (Fig. 5). Stimulation was applied to elicit activation of the phantom hand regions to correspond with the appropriate movement. There was higher activation in the central and centro-parietal regions during the Stim-Move condition compared to the Pre-Stim condition (Fig. 7C, I). In the Post-Stim condition, the effect of the stimulation persisted and changes in neural activity were observed in the central and the centro-parietal regions (Fig. 7D, J). We also compared the alpha power in individual central and centro-parietal electrodes across the conditions in both amputees (Fig. 7E-F, K-L). One-way ANOVA followed by post-hoc analysis was performed for each of the electrodes. In both participants, significant increases in the relative alpha power was observed for phantom hand movement during the Stim-Move and Post-Stim conditions compared to the Pre-Stim condition. Interestingly, the largest change in relative alpha power in the neural signal occurred in the ipsilateral hemisphere of A02, relative to the phantom hand. A03’s neural activity showed changes in both the contralateral and ipsilateral hemispheres.

**Fig. 7.**
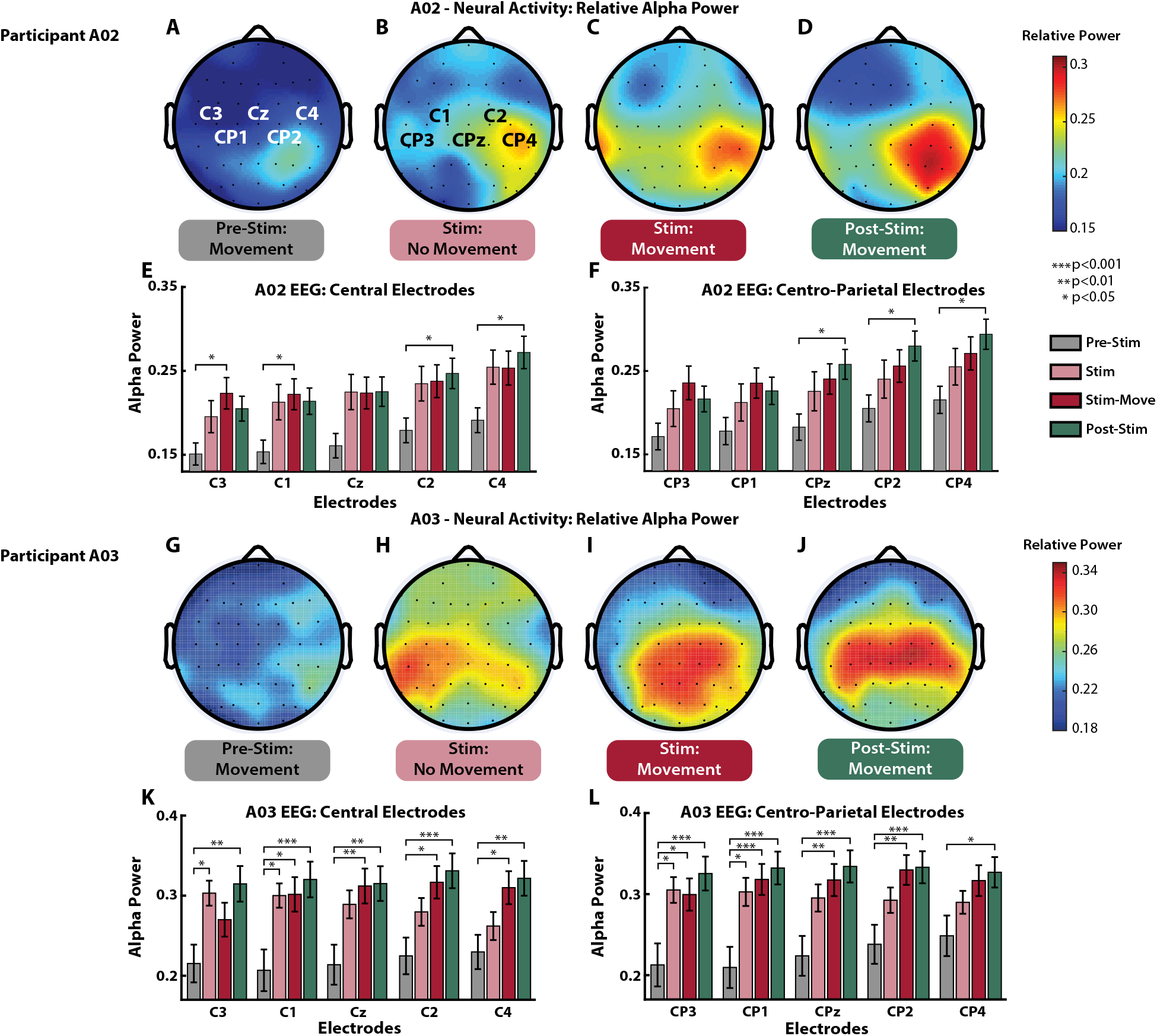
Neural activity in sensorimotor regions. Participants A02 and A03 received visual cues and performed the corresponding hand movements during tTENS (Fig. S6). Sensory stimulation was given through tTENS and recordings for each condition (Pre-Stim, Stim, Stim-Move, Post-Stim) were performed in order with <10 min between conditions. **(A-D)** A02’s relative alpha power neural activation maps for movements before any sensory stimulation (Pre-Stim), stimulation with no phantom hand movements (Stim), movements with sensory stimulation (Stim-Move), and phantom hand movements (Post-Stim). The movements were hand open and close. Each grip corresponded to stimulation of different regions of the phantom hand (Fig. S7). **(E-F)** A02’s relative alpha power in the central and centro-parietal electrodes, respectively. For all conditions n = 60. **(G-J)** A03’s relative alpha power neural activation maps across the various conditions. The movements were tripod, index point, and wrist flexion. Each grip corresponded to stimulation of different regions of the phantom hand (Fig. S7). For Pre-Stim n=30 and for all other conditions n=60. **(K-L)** A03’s relative alpha power in the central and centro-parietal electrodes, respectively. There were noticeable changes in general neural (Fig. S8) and alpha band activity as a result of stimulation and these changes persisted during the Post-Stim condition.

## 4. Discussion

### 4.1. Sensory stimulation improves perception

Our results show activation and enhanced perception of the phantom limb from tTENS (Fig. 2). Typically this technique is used to provide tactile sensations to the phantom hand [10, 12, 26]. Interestingly, the heightened sense of the phantom limb also seems to relate to changes in muscle activity during movements. The amputees felt as if the sensations were more or less natural, reported primarily as being a pressure or buzzing, and originating from their phantom hand. It is unclear if A01’s thermal sensation was a result of dominant thermal-specific afferents or a residual effect of the recent amputation. A01 described his phantom hand as a “foggy” and buzzing sensation as a result of the recent amputation. In the survey, the amputees indicated stronger perception of the phantom hand as a result of stimulation, which enabled a greater ability to move their phantom hand despite its absence (Fig. 3).

Remarkably, over 2 years the stimulation sites and perceived activated regions in the phantom hand remained relatively stable for subject A03 (Fig. 5A-B, and Fig. S3). Despite an amputation over 7 years prior to the study, the sensory nerves in the residual limb still provided meaningful sensations of touch back to the user indicating cortical representation of the phantom hand as well as intact neural pathways. Although there are slight differences in activated sensory maps each day, the activated phantom regions themselves did not migrate and retained structural similarity over time (Fig. 5B), suggesting no major changes in the area of perceived activation. The fact that the sensory maps did not significantly change suggests that phantom limb representation remains many years after injury even without constant sensory stimulation.

### 4.2. Phantom limb perception improves movement decoding

Our results suggest that the internal sensorimotor pathway is affected by stimulation and enhanced phantom limb perception (Fig. 4). A crucial aspect of controlling the phantom hand, and in turn a prosthesis, is the internal perception of the phantom limb. Sensory feedback can be used to convey tactile information back to amputees [6–8, 12, 14–16]; however, we show that phantom hand perception is fundamentally linked to motor performance even in the absence of object manipulation.

Our results suggest that sensory stimulation influences real-time myoelectric pattern recognition and that enhanced phantom perception temporarily improves movement decoding, regardless of experience level (Fig. 5 and 6). By working closely with patients A02 and A03, we identified the most relevant regions of the phantom hand to enhance perception during certain movements. A03 believed it would be difficult to achieve reliable control of more than 9 movement classes as a non-TMR transhumeral prosthesis user. To see how much improvement was possible due to strengthening the internal sensorimotor control loop of the amputee, we expanded the number of classes to 14 (Fig. 5C-F).

Participant A03 had previous experience with myoelectric pattern recognition and did not show significant improvement as a result of additional training over Phase I of the long-term study (Fig. 5G); however, there were significant improvements during the sessions with sensory stimulation to the phantom hand (Phase II, Fig. 5H). These results indicate that the heightened sense of the phantom hand immediately strengthens the sensorimotor loop of the amputee, but this improvement does not extend across days if the stimulation does not persist. Periodic sensory reinforcement provides short-term benefit, but the ability to perform the movements was similar during both Phases I and III, with a slight upward trend throughout the study, indicating no long-term impact and likely a result of continued prosthesis use by A03 (Fig. 5G). Long-term improvements may be realized through continued sensory reinforcement over a longer period.

The link between phantom perception and control is evident from the results, and it is supported by prior work that shows motor cortex excitability can increase with sensory activation [35]. According to participant A03, enhanced sensation of the phantom hand from tTENS can take up to several hours to subside after which the phantom hand perception returns to the baseline state. The temporary influence of tTENS aligns well with our observed improvements in motor control being limited to within a single day. The subsiding effect of the sensory stimulation and movement decoding improvements limited to a single day further supports the idea that recurring sensory stimulation sessions could help create more permanent improvement. It is possible that a more targeted prosthesis training paradigm, making use of combined phantom motor control and sensory stimulation, would lead to long-term improvements in prosthesis performance. It should be noted that we did not compare non-phantom hand sensory stimulation conditions to investigate if the improved movement decoding effect is also present during other types of sensory activation to the body. Regardless, the results suggest a relationship between phantom perception and movement decoding, which may prove valuable for improving prosthesis control. We also showed a slight improvement in task completion time after sensory activation of the phantom hand during prosthesis control in an object movement task (Fig. 6E), which further supports the idea of phantom hand perception playing an important role in functional prosthesis control.

### 4.3. Sensory stimulation activates sensorimotor regions

Our neurological studies based on EEG activation present evidence for sensory-motor integration in amputees. The central and centro-parietal electrodes cover the primary motor and somatosensory cortices, which are areas known to be activated during sensory processing and motor-related tasks [36, 37]. In stroke patients, median nerve activation using TENS is also known to increase motor cortex excitability and motor function [35]. Based on our results, we inferred that tTENS does not just act as a tactile stimulus but also improves the perceived control of the phantom hand by the amputee. This inference is supported by the improvements in movement decoding (Fig. 4, 5, 6) and aligns with previous results suggesting the role of central and parietal EEG activation in phantom limb vividness [38]. Previous studies have also demonstrated central and somatosensory cortical region EEG activation [10, 12, 39] and enhanced connectivity [40] from noninvasive sensory feedback; however, here we also observed that the sensory stimulation effect persisted during the Post-Stim condition. It should be noted that the above observations were made for all the stimulation sites (median, ulnar and radial). We did not observe significant differences between them owing to the lower spatial resolution of EEG. Nevertheless, EEG-based classification of A03’s stimulation sites was possible with relatively high accuracy (Fig. S6).

We believe that during the Post-Stim condition the activation of the primary somatosensory cortex showed tactile working memory aiding the amputee in better perception of the phantom hand movements even without the feedback stimulus. Prior work showed that the primary sensory cortex (both contralateral and ipsilateral) acts as a center for online sensory processing as well as a transient storage site for tactile information [41, 42]. It’s possible that sensory stimulation to the phantom hand aids the amputees to make movement because the tactile working memory plays a valuable role in aiding the movement and perception both during and after the sensory stimulation.

Interestingly, the most significant neural changes in A02 occurred in the ipsilateral hemisphere during tTENS (Fig. 7B-D). While the traditional notion is that sensorimotor activity is in the contralateral hemisphere, previous work showed decoding motor commands from ipsilateral brain activity in the sensorimotor region [43]. The potential roles of ipsilateral activity suggested in the past include contributing to finger representation and voluntary execution of a movement [44] and maintaining an efference copy and muscle posture of the ipsilateral limb [45]. Another possibility of stronger ipsilateral activation could be to the absence of contralateral inhibition due to cortical adjustments after injury and amputation [46]. Because we know both hemispheres are used for tactile information storage, it’s likely that short-term tactile working memory was utilized [41, 42].

Furthermore, the laterality differences observed in amputees A02 and A03 could be the result of time since amputation, individual variability in amputation, and experiences in tTENS and myoelectric control. Participant A02 had 15 years between paralysis from nerve injury and elective amputation (Table S1). Though the extent of peripheral sensory input loss effects in cortical behavior is not completely understood, cortical functional organizational differences could also contribute to the differences observed in A02 and A03. Participant A03 received an extended period of sensory mapping (Fig. 5A, B). Studying the long-term cortical effects of sensory stimulation can offer more insights to cortical behavior when sensory information is reintroduced to amputees. In targeted muscle and sensory reinnervation (TMSR) recipients, the strength in activation of primary motor and somatosensory regions shows similarity with that in intact limb controls [47].

Despite disrupted sensorimotor pathways after limb amputation, there is bilateral activation during electrical sensory stimulation [39] and phantom movements [48]. Such cortical plasticity mechanisms are not completely understood, but deeper insight could be obtained from studying the causal interactions between the two hemispheres focusing on the sensorimotor loop. Our observations on enhanced phantom perception influencing control and neural activation support the idea that phantom hand representation in the cortex persists after amputation [22, 24]. More research is needed to develop methods sustain movement decoding improvement beyond a single day; however, our EEG results offer insight on the role of sensory feedback in phantom hand perception and control.

## 5. Conclusion

We show that improving perception of the phantom hand in amputees can improve the ability to produce and control phantom hand movements. This improvement in phantom hand control can be captured and decoded through surface EMG. Sensory stimulation of the phantom hand appears to provide short-term (within a single day) improvements in movement decoding. Nerve stimulation can provide tactile feedback for amputees, but here we show that sensory stimulation of the phantom hand is also fundamentally linked to phantom hand control. Through EEG signals, we confirm that the sensory activation of the phantom hand influences relevant neural motor activity. Interestingly, the enhanced neural motor activity persists even after sensory stimulation is removed, which helps explain the movement decoding improvements after short phantom hand sensory stimulation sessions. When tracked over 2 years, we saw that the sensory regions of the phantom hand did not change, which demonstrates long-term stability in amputee sensory maps even many years after amputation. Movement decoding performance over 1 year did not substantially change; however, performance was affected on the same day as targeted phantom hand sensory stimulation. Our findings offer insight on how phantom hand perception can be modulate through sensory activation for improving motor control, prosthesis function, and rehabilitation after amputation.

## Data Availability

All data needed to evaluate the claims are contained within the manuscript and supplemental material.

## Acknowledgments

This work was supported by Space@Hopkins, the National Institutes of Health (T32EB003383), the National Science Foundation (1849417), and the JHU/APL postdoctoral fellowship. The VIE was developed at JHU/APL under the Revolutionizing Prosthetics program (Defense Advanced Research Projects Agency, N66001-10-C-4056).

## Author contributions

L.E.O., K.D., M.A.H., G.M.L., and C.L.H. developed hardware and software for the experiments. L.E.O., M.A.H., K.D., and M.M.I. conducted experiments. L.E.O. and N.V.T. designed the experiments. L.E.O., M.A.H., R.B., K.D., M.M.I., A.D., Z.T., A.B., and N.V.T. analyzed the data. N.V.T. supervised all experiments, data analysis, and interpretation of the results. All authors contributed to writing the paper.

## ORCID iD

Luke E. Osborn: 0000-0003-2985-6294

Keqin Ding: 0000-0002-0680-673X

Rohit Bose: 0000-0003-3966-4464

Mark M. Iskarous: 0000-0003-4208-3943

Andrei Dragomir: 0000-0003-2815-6958

Zied Tayeb: 0000-0003-3257-0211

Gorden Cheng: 0000-0003-0770-8717

Robert S. Armiger: 0000-0002-3437-1884

Anastasios Bezerianos: 0000-0002-8199-6000

Matthew S. Fifer: 0000-0003-3425-7552

Nitish V. Thakor: 0000-0002-9981-9395

